# Impact of COVID-19 pandemic on the incidence of suicidal behaviors: a retrospective analysis of integrated electronic health records in a 7.5-million population

**DOI:** 10.1101/2022.09.26.22280233

**Authors:** Damià Valero-Bover, Marc Fradera, Gerard Carot-Sans, Isabel Parra, Jordi Piera, Caridad Pontes, Diego Palao

## Abstract

The COVID-19 pandemic has caused a remarkable psychological overwhelming and an increase of stressors that may trigger suicidal behaviors. However, its impact on the rate of suicidal behaviors has been poorly reported. We conducted a population-based retrospective analysis of all suicidal behaviors attended in healthcare centers of Catalonia (North-East Spain; 7.5-million inhabitants) between January 2017 and June 2022. We retrieved data from the episode, including an assessment of suicide risk and the individual’s socioeconomic and clinical characteristics. Data were summarized yearly and for the periods before and after the onset of the COVID-19 pandemic in Spain in March 2020. The analysis included 26,458 episodes of suicidal behavior (21,920 individuals); of them, 16,414 (62.0%) were suicide attempts. The monthly moving average ranged between 300 and 400 episodes until July 2020, and progressively increased to over 600 episodes monthly. In the post-pandemic period, suicidal ideation increased at the expense of suicidal attempts. Cases showed a lower suicide risk; the percentage of females and younger individuals increased, whereas the prevalence of classical risk factors, such as living alone or lacking a family network and a history of psychiatric diagnosis, decreased. In summary, suicidal behaviors have increased during the COVID-19 pandemic, with more episodes of suicidal ideations without attempt and younger and lower risk profiles.

## 1. Introduction

The severe acute respiratory syndrome coronavirus 2 (SARS-CoV-2) spread rapidly from China, where the first cases were identified in December 2019, to all around the globe, leading to an unprecedented global health crisis. Aside from the direct effects of the coronavirus disease 2019 (COVID-19), the pandemic has had other important impacts derived from the measures dictated for limiting the SARS-CoV-2 spread. These measures, primarily based on quarantines (either general or targeting cases and contacts), social distancing, and banning of gatherings [1], have altered the social interactions, with potential impact to mental health, particularly among individuals with previous psychiatric pathologies [2–5]. Aside from the stressors directly derived from social distancing, other factors such as uncertainty associated with the pandemic scenario, impossibility of accompanying loved ones during their hospital stay or end-of-life stage, or financial crises (including unemployment) experienced in many countries have contributed to psychological overwhelming in many cases [6]. Likewise, the persistence of the health crisis, with alternating periods of normal live and social restrictions due to successive waves of the COVID-19 has caused learned helplessness behaviors, increasing the risk of depressive disorders [7]. Irrespective of the cause of mental health decline, various authors have reported increasing use of mental health services during the pandemic [3,4].

Overwhelming, experiencing disaster, and sense of isolation are among the triggers of suicidal behaviors, particularly among individuals with underlying mental disorders, such as depression, schizophrenia, or alcohol abuse, among others [8]. In 2019, before the onset of the COVID-19 pandemic, the number of deaths due to suicide reported by the WHO amounted to over 700,000 cases worldwide, with suicide being the fourth leading cause of death among 15-to-29 year olds [9]. Although the report showed a decline in suicide rates in Europe between 2000 and 2019, the impact of the COVID-19 pandemic on mental health is likely to revert this trend, and figures in some countries suggest that the death toll due to suicide could be even greater than that directly attributed to SARS-CoV-2 infection [10,11].

To date, various authors have reported changes in patterns and lethality of suicidal behaviors associated with the COVID-19 pandemic [12–15]. However, the lack of integrated health information in many countries limit the number of population-based analysis of the shift in suicide rates before and after the COVID-19 pandemic. Likewise, little is known on whether the profile of individuals who attempt suicide before and after the pandemic onset is changing. In this retrospective analysis of a 7.5 million population, we have investigated the changes in suicidal behaviors and individual profile before and during the first two years of the COVID-19 pandemic.

## 2. Materials and Methods

### 2.1. Study design and setting

This was a retrospective analysis of electronic health records of Catalonia, a 7.5-million people area in the North-East Spain. The Catalan Health Service (CHS) provides public, universal healthcare to the entire population of Catalonia. Although information systems are fragmented into various datasets of hospital, primary care, and mental health, among others, the unique identification number used for insuring purposes allows integration of information across the entire system.

In 2014, the CHS launched the Suicide Risk Code, a secondary prevention program aimed at reducing mortality due to suicide, increase survival among individuals attended because of suicidal behaviors, and prevent repeated suicidal attempts in patients with risk factors [16]. One of the components of the Suicide Risk Code was a specific registry for recording all cases of suicidal behavior attended in CHS health centers and collect information regarding the episode, key clinical and social conditions, and care pathway followed after the episode. The Suicide Risk Code was progressively implemented until covering the entire territory of Catalonia by 2016. Since then, all individuals with a suspected risk of suicide visited in any healthcare center of the CHS are referred to the closest psychiatry ward for a comprehensive assessment, and registered into the Suicide Risk Code dataset, regardless of the presence or not of a suicide attempt. To prevent biases due to incomplete implementation of the program, for this analysis, we screened the Suicide Risk Code dataset for episodes occurred from January 01, 2017, to the end of the investigated period in June 30, 2022.

The study protocol was approved by the independent ethics committee of the Consortium Corporació Sanitària Parc Taulí of Sabadell, which waived the collection of informed consent for secondary use of health data collected during routine care. The study was conducted according to the General Data Protection Regulation 2016/679 on data protection and privacy for all individuals within the European Union and the local regulatory framework regarding data protection.

### 2.2. Variables and data sources

The characteristics of the suicidal behavior episode and risk factors recorded at the time of the episode were retrieved from the Suicide Risk Code dataset. The type of suicidal behavior episode was classified as either active suicidal ideation without attempt or suicide attempt, defined as engaging in self-directed injury with specific intent to die. In patients with suicide attempt, the method used in the attempt was recorded. The following variables were recorded during the visit triggering the entry in the Suicide Risk Code registry: family history of suicide, severe or painful systemic disease, abuse of alcohol or drugs, recent economic or personal stressors, lack of family or social network, living alone, and recent hopelessness feelings. We also collected the suicide risk level, rated based on the six-item suicidality module of the Mini International Neuropsychiatric Interview (MINI) [17]. The suicidality score ranges from 0 to 33, and allows grouping the risk into low (0 – 5 points), moderate (6 – 9 points), and high (≥ 10 points).

The CHS identification number was used to retrieve demographic characteristics (i.e., age and sex) and psychiatric disorders diagnosed before or up to one moth following the suicidal behavior episode: schizophrenia and schizoaffective disorders, eating disorders, anxiety, depression (including recurrent or persistent mood (affective) disorders), disorders of personality and behavior, sleep disorders, Alzheimer disease, and psychoactive substance use. The International Classification of Diseases version 10 – Clinical Modification (ICD-10-CM) codes are listed in Table S1 of the Supplementary File 1.

Finally, we collected the socioeconomic status based on the pharmaceutical co-payment classification of the CHS, which stratifies the population into four socioeconomic groups based on the pharmaceutical co-payment: very low (i.e., individuals perceiving a minimum integration income, unemployment allowance, unemployment benefit, those on leave for work-related accident or professional disease, persons with severe disability, and other highly vulnerable groups), low (annual income < € 18,000), moderate (annual income € 18,000 to € 100,000), and high (annual income > € 100,000). During the 2020 – 2021 period, the criteria for very low and low groups changed to widen coverage of the very low group; therefore, for the purpose of this work, we grouped these two categories.

### 2.3. Analysis

The progression of suicidal behavior episodes throughout the investigated period was plotted as the monthly incidence (observed and moving average). The moving average considered a 12-month symmetric window with equal weights. The seasonal component was computed by averaging, for each time unit, over all periods, whereas the error component was determined by removing trend and seasonal components from the original time series. The monthly incidence was determined for the entire sample and for age, sex, and socioeconomic groups of interest. Data on Catalan population was retrieved from the Statistical Institute of Catalonia for incidence estimate [18]. The characteristics of the episode and clinical and demographic profile of individuals were described with the frequency and percentage over available data. Continuous variables were described using the mean and standard deviation (SD). The episode characteristics and individual profile were summarized for each natural year of the investigated period. Additionally, we estimated the percentage of each risk factor and episode characteristic before and after the onset of the COVID-19 pandemic in Catalonia, in March 2020, and considering natural months as units. The analysis was descriptive and no hypothesis tests were performed. All analyses were conducted in R version 4.0.4 [19]; plots were built using package ggplot version 3.3.3 [20].

## 3. Results

### 3.1. Incidence of suicidal behavior

Between January 2017 and June 2022, the Suicide Risk Code registered 26,458 episodes of suicidal behavior in 21,920 individuals. The monthly rate of suicidal behavior events dropped extremely during the two months of nationwide lockdown and the overall trend remarkably increased afterwards (Figure 1a). Overall, the rate of suicidal behaviors per 100,000 inhabitants per month ranged from 3.06 to 6.17 before the pandemic onset (2017 – 2019, both included) and 7.43 to 9.42 within the last six months of the observation period (January – June 2022). The incidence increase was more pervasive among females (Figure 1a) and minors (Figure 1b). The subgroup analysis according to gender and age confirmed females under 18 years old as the primary contributors to the overall increase in suicidal behaviors in the general population (Figure 1c). The analysis according to the socioeconomic status showed increases in all groups, with an incidence persistently higher in the low socioeconomic group (Figure 1d). The higher incidence increase among female and minors was consistent across all socioeconomic groups (Figure S2, Supplementary file 1).

**Figure 1.**
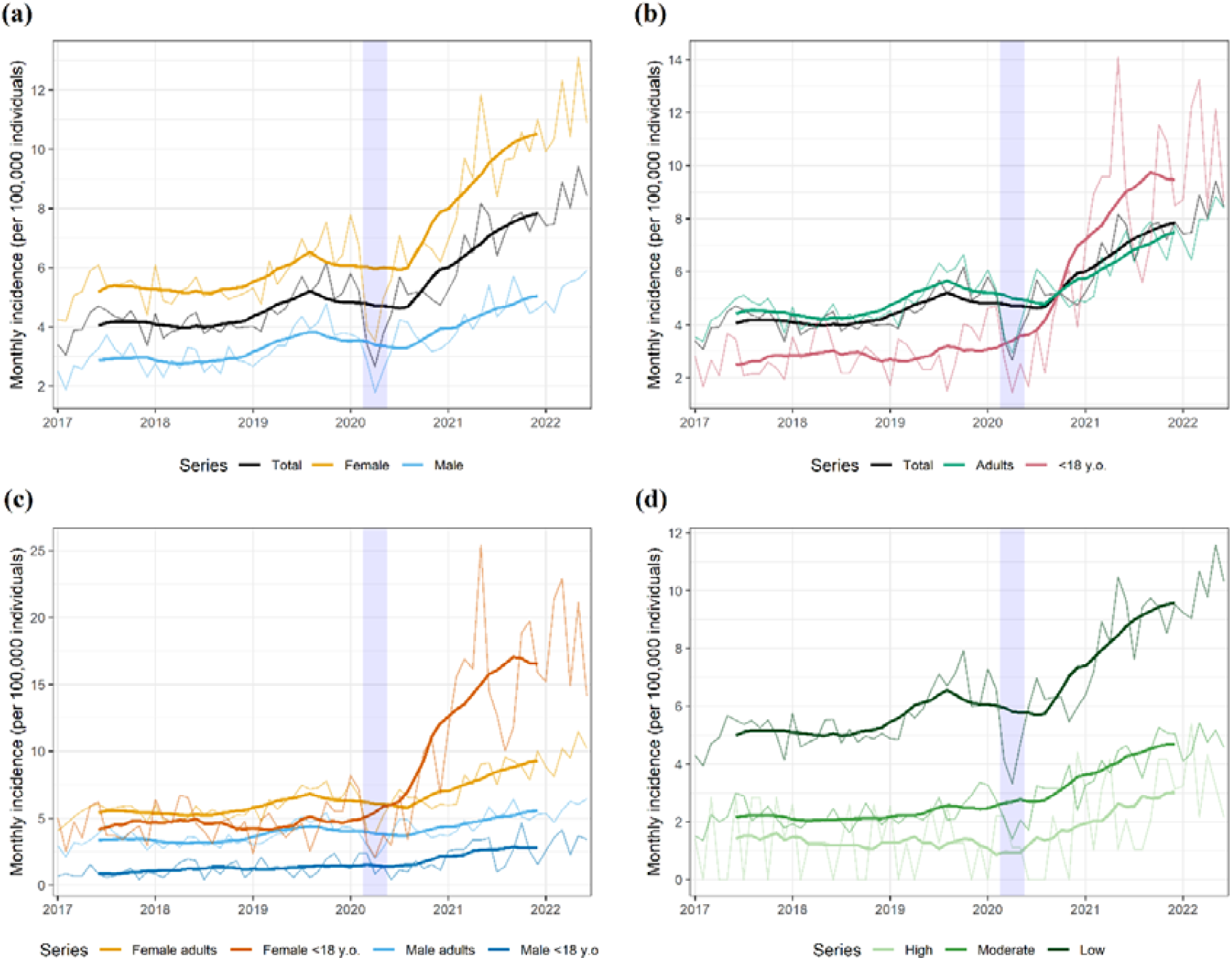
Monthly rate of suicidal behavior episodes for the overall population and stratified according to (a) gender, (b) age group, (c) gender and age group, and (d) socioeconomic status. The shaded area represents the nation-wide lockdown period, which was dictated on March 13, 2020, started a progressive easing on May 10, 2020, and was definitely lifted on June 20, 2020. The seasonal and random components of the observed rate are presented in Figure S1 (Supplementary file 1).

### 3.2. Risk factors and characteristics of suicidal behaviors

Table 1 summarizes the characteristics of the suicidal behavior episodes through the investigated period. Compared with the three years before the onset of the COVID-19 pandemic, episodes reported during the pandemic were less frequently an attempt, and individuals showed a lower suicidal risk in the MINI assessment at the time of the attempt. The mean (SD) MINI scores were 13.5 (8.9), 12.9 (8.4), and 12.0 (8.3) for 2017, 2018, and 2019, respectively, and 11.6 (8.4), 11.1 (8.1) and 11.1 (8.0) for 2020, 2021, and first semester of 2022.

**Table 1.**
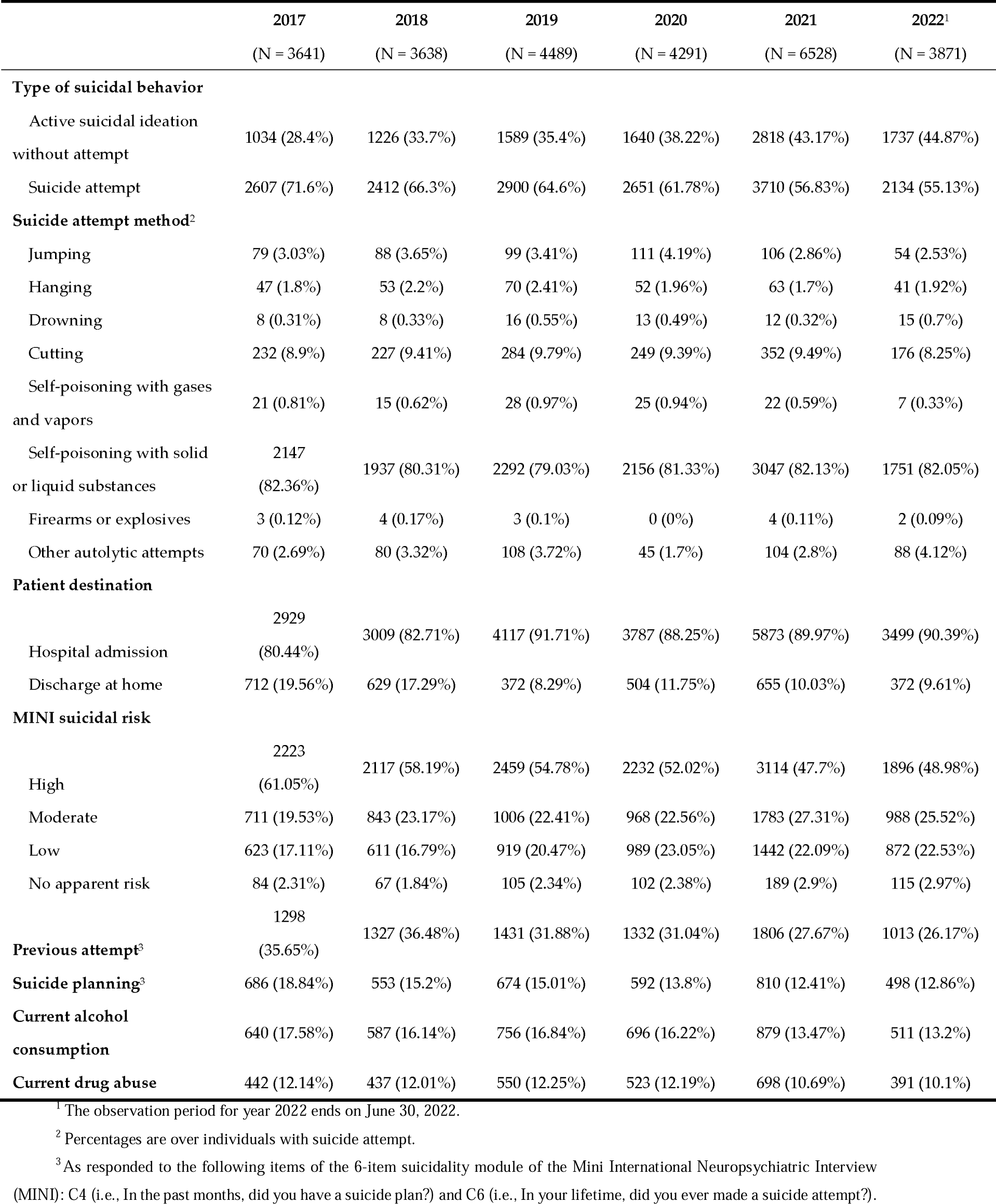
Characteristics of the episode through the investigated period. Results are presented as no. and percentage of episodes.

|  | 2017<br>(N = 3641) | 2018<br>(N = 3638) | 2019<br>(N = 4489) | 2020<br>(N = 4291) | 2021<br>(N = 6528) | 2022<sup>1</sup><br>(N = 3871) |
| --- | --- | --- | --- | --- | --- | --- |
| **Type of suicidal behavior** |  |  |  |  |  |  |
| Active suicidal ideation without attempt | 1034 (28.4%) | 1226 (33.7%) | 1589 (35.4%) | 1640 (38.22%) | 2818 (43.17%) | 1737 (44.87%) |
| Suicide attempt | 2607 (71.6%) | 2412 (66.3%) | 2900 (64.6%) | 2651 (61.78%) | 3710 (56.83%) | 2134 (55.13%) |
| **Suicide attempt method<sup>2</sup>** |  |  |  |  |  |  |
| Jumping | 79 (3.03%) | 88 (3.65%) | 99 (3.41%) | 111 (4.19%) | 106 (2.86%) | 54 (2.53%) |
| Hanging | 47 (1.8%) | 53 (2.2%) | 70 (2.41%) | 52 (1.96%) | 63 (1.7%) | 41 (1.92%) |
| Drowning | 8 (0.31%) | 8 (0.33%) | 16 (0.55%) | 13 (0.49%) | 12 (0.32%) | 15 (0.7%) |
| Cutting | 232 (8.9%) | 227 (9.41%) | 284 (9.79%) | 249 (9.39%) | 352 (9.49%) | 176 (8.25%) |
| Self-poisoning with gases and vapors | 21 (0.81%) | 15 (0.62%) | 28 (0.97%) | 25 (0.94%) | 22 (0.59%) | 7 (0.33%) |
| Self-poisoning with solid or liquid substances | 2147 (82.36%) | 1937 (80.31%) | 2292 (79.03%) | 2156 (81.33%) | 3047 (82.13%) | 1751 (82.05%) |
| Firearms or explosives | 3 (0.12%) | 4 (0.17%) | 3 (0.1%) | 0 (0%) | 4 (0.11%) | 2 (0.09%) |
| Other autolytic attempts | 70 (2.69%) | 80 (3.32%) | 108 (3.72%) | 45 (1.7%) | 104 (2.8%) | 88 (4.12%) |
| **Patient destination** |  |  |  |  |  |  |
| Hospital admission | 2929 (80.44%) | 3009 (82.71%) | 4117 (91.71%) | 3787 (88.25%) | 5873 (89.97%) | 3499 (90.39%) |
| Discharge at home | 712 (19.56%) | 629 (17.29%) | 372 (8.29%) | 504 (11.75%) | 655 (10.03%) | 372 (9.61%) |
| **MINI suicidal risk** |  |  |  |  |  |  |
| High | 2223 (61.05%) | 2117 (58.19%) | 2459 (54.78%) | 2232 (52.02%) | 3114 (47.7%) | 1896 (48.98%) |
| Moderate | 711 (19.53%) | 843 (23.17%) | 1006 (22.41%) | 968 (22.56%) | 1783 (27.31%) | 988 (25.52%) |
| Low | 623 (17.11%) | 611 (16.79%) | 919 (20.47%) | 989 (23.05%) | 1442 (22.09%) | 872 (22.53%) |
| No apparent risk | 84 (2.31%) | 67 (1.84%) | 105 (2.34%) | 102 (2.38%) | 189 (2.9%) | 115 (2.97%) |
| Previous attempt<sup>3</sup> | 1298 (35.65%) | 1327 (36.48%) | 1431 (31.88%) | 1332 (31.04%) | 1806 (27.67%) | 1013 (26.17%) |
| Suicide planning<sup>3</sup> | 686 (18.84%) | 553 (15.2%) | 674 (15.01%) | 592 (13.8%) | 810 (12.41%) | 498 (12.86%) |
| Current alcohol consumption | 640 (17.58%) | 587 (16.14%) | 756 (16.84%) | 696 (16.22%) | 879 (13.47%) | 511 (13.2%) |
| Current drug abuse | 442 (12.14%) | 437 (12.01%) | 550 (12.25%) | 523 (12.19%) | 698 (10.69%) | 391 (10.1%) |

<sup>1</sup> The observation period for year 2022 ends on June 30, 2022.
<sup>2</sup> Percentages are over individuals with suicide attempt.
<sup>3</sup> As responded to the following items of the 6-item suicidality module of the Mini International Neuropsychiatric Interview (MINI): C4 (i.e., In the past months, did you have a suicide plan?) and C6 (i.e., In your lifetime, did you ever made a suicide attempt?).

The analysis of the underlying social and clinical characteristics of the cases through the investigated period revealed an increase in the percentage of younger individuals and females in the years following the pandemic onset (Table 2). Also, individuals with suicidal behaviors within the years following the pandemic onset were less likely to present with classical risk factors for suicide attempt, such as living alone, lacking social or family support, or having an underlying mental disorder.

**Table 2.**
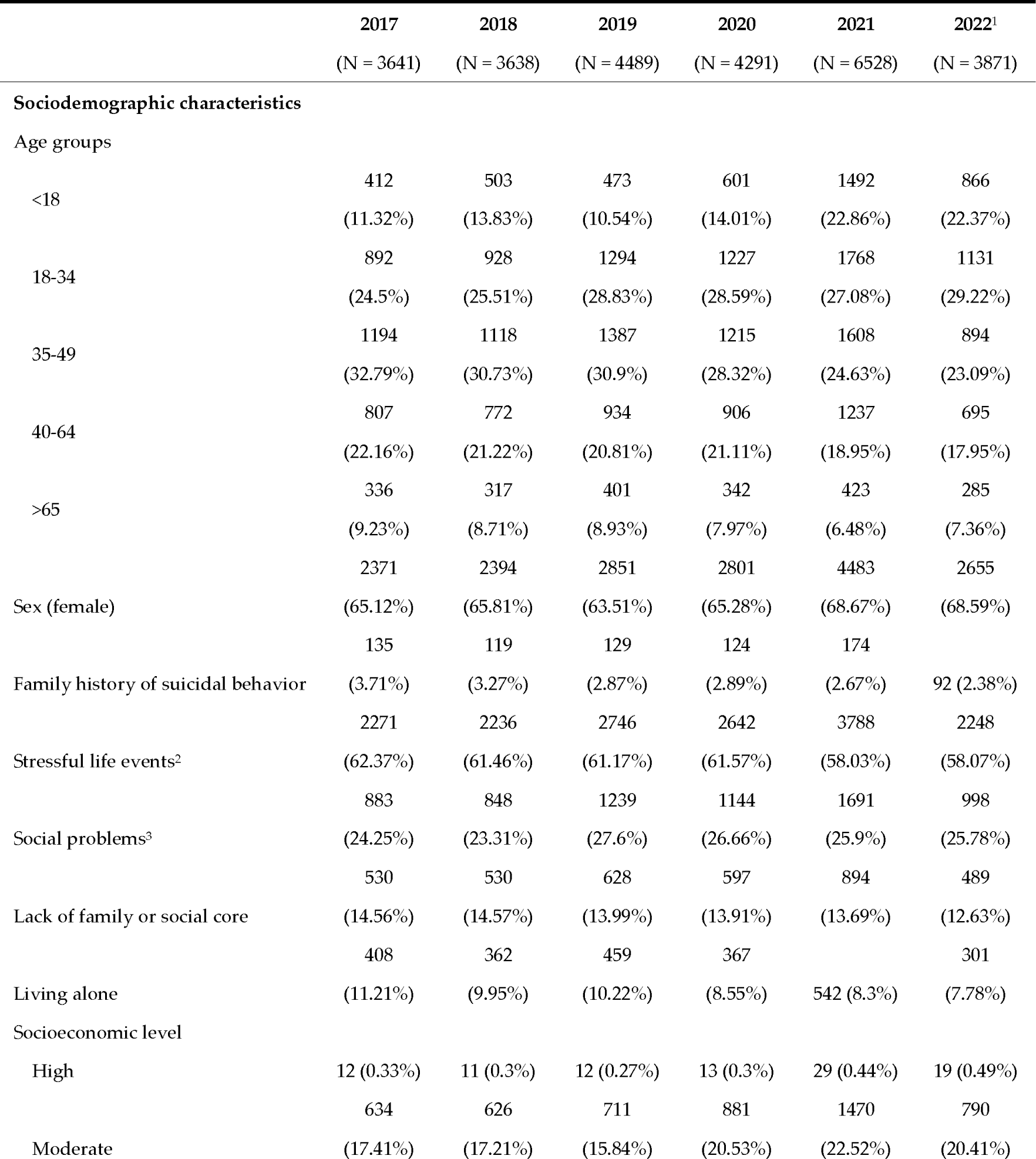
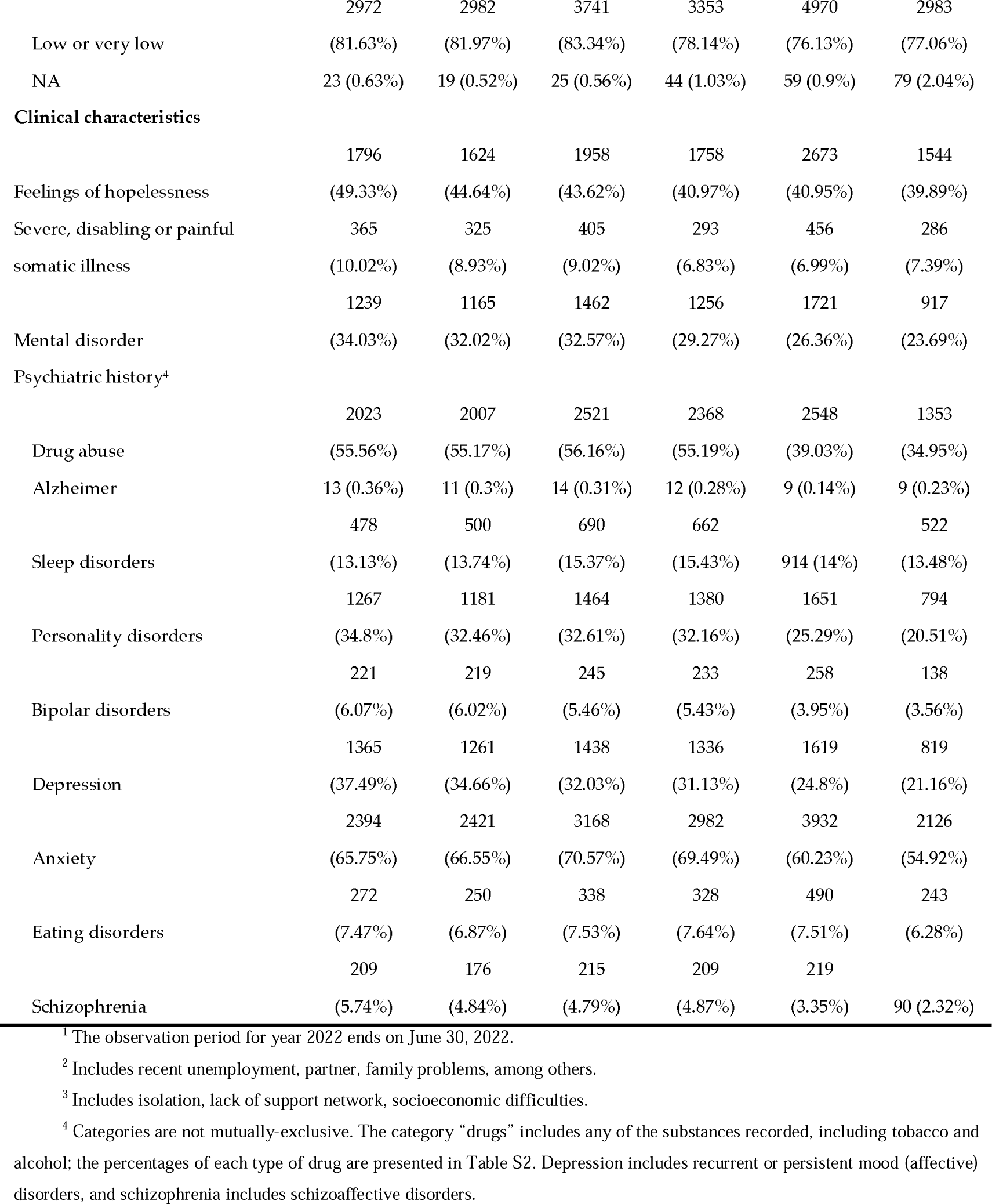
Sociodemographic and clinical characteristics of individuals with suicidal behaviors within the investigated period. Results are presented as no. and percentage of individuals.

The average values before and after the pandemic onset confirmed the trends observed in the yearly analysis (Figure 2). In both periods, self-poisoning with liquid or solid substances remained the leading method among those with attempt, with no remarkable changes observed regarding the frequency of each method between periods (Figure 2a). However, after the pandemic onset, suicidal behavior episodes were more frequently ideations without a suicide attempt. The mean number of suicidal behavior episodes per case was similar before (1.3 [SD 0.9]) and after (1.4 [1.0]) the pandemic onset.

**Figure 2.**
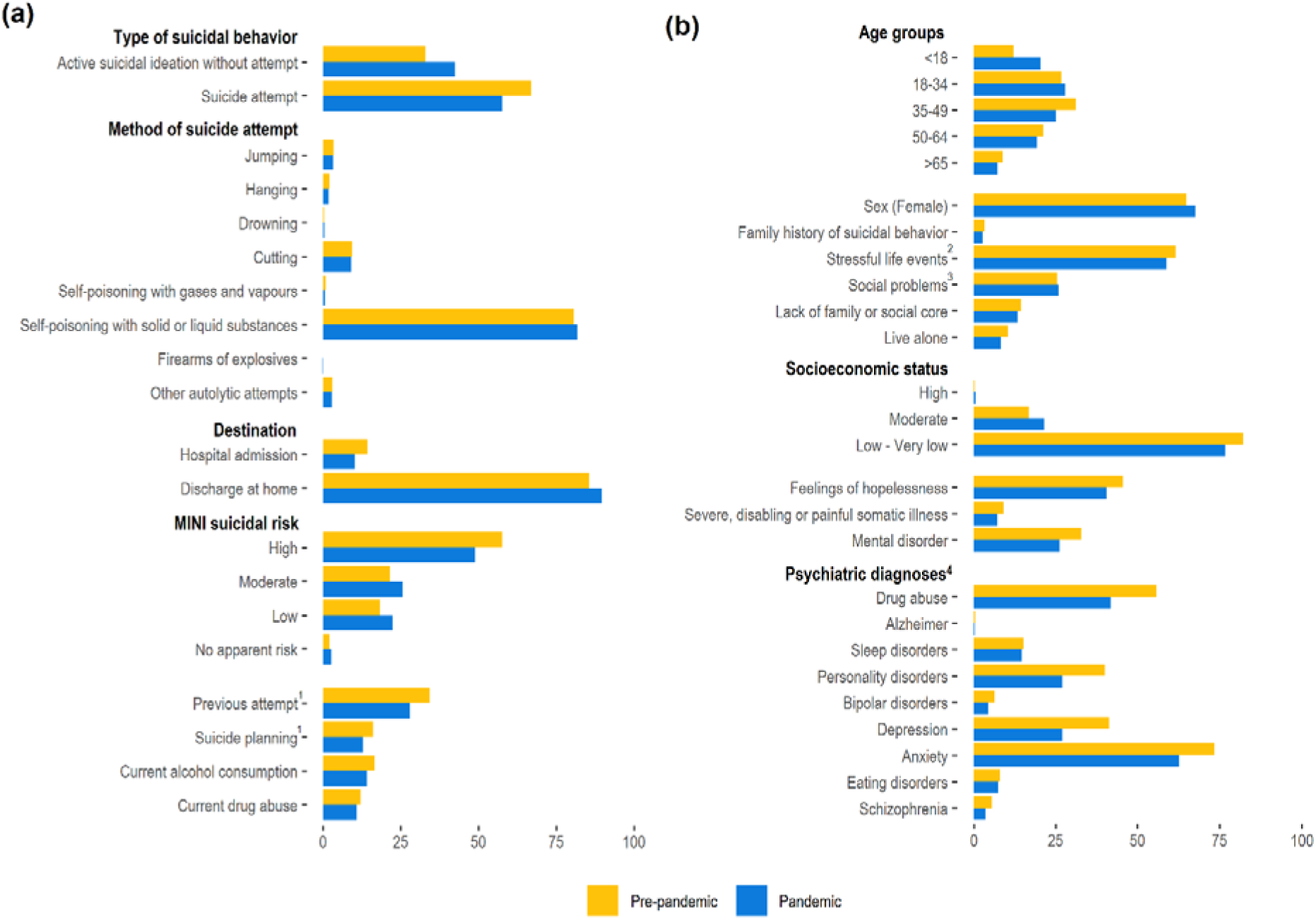
Characteristics of the episodes (a) and individuals (b) with suicidal behaviors. Values correspond to the average for the pre-pandemic period (i.e., January 01, 2017 to February 29, 2020) and pandemic period (i.e., March 01, 2020 to June 30, 2022). ^1^ As responded to the following items of the 6-item suicidality module of the Mini International Neuropsychiatric Interview (MINI): C4 (i.e., In the past months, did you have a suicide plan?) and C6 (i.e., In your lifetime, did you ever made a suicide attempt?). ^2^ Includes recent unemployment, partner, family problems, among others. ^3^ Includes isolation, lack of support network, socioeconomic difficulties. ^4^ Categories are not mutually-exclusive. The category “drug abuse” includes any of the substances recorded (the percentages for each type of drug are listed in Figure S3). Depression includes recurrent or persistent mood (affective) disorders, and schizophrenia includes schizoaffective disorders.

The most remarkable change in the individual profile was the lower frequency of mental disorders after the pandemic onset, evidenced in all psychiatric diagnoses, particularly in schizophrenia and schizoaffective disorders (Figure 2b). Depression, anxiety, and drug use were consistently the most prevalent mental health disorders.

## 4. Discussion

In this population-based, retrospective analysis of suicidal behaviors among a 7.5-million population, we found that the COVID-19 crisis led to a remarkable increase in the rate of suicidal behavior episodes, primarily explained by an incidence increase among young females. The turning point in the trend of suicidal behavior was pervasive after June 2020, four months after the onset of the outbreak in our country. We also found changes in the profile of individuals with suicidal behaviors, who presented with a lower prevalence of classical risk factors for suicide and suicidal behaviors, such as alcohol/drug abuse or underlying psychiatric disorders.

The delayed turning point observed in suicidal behaviors is consistent with retrospective analyses conducted early during the pandemic. Reif-Leonhard et al. found a lower rate of suicide attempts reported at the Frankfurt program to prevent suicides in 2020 compared to 2019 [13]. Similarly, the demand for the suicide prevention helpline in the Netherlands decreased within the first months of the pandemic and started an increasing trend approximately four months after the first case in the country [12]. Finally, Yoshioka et al., observed that the decreasing trend in suicides experienced in Japan during the 2016-2019 period was reverted in the overall 2020-2021 period, although a delay of several months in the rough observed cases was reported [15]. Interestingly, the strict lockdown period, an important stressor associated with an increased number of anxiety episodes and psychiatric consultations in our area [7,21], was followed by a drop in suicidal behavior events in our analysis. This phenomenon, observed also in a city-based study in our area [22], is in line with the decreasing demand for the suicide prevention helpline observed in a nation-wide analysis in the Netherlands right after the general lockdown [12]; of note, a partial lockdown dictated in that country eight months after was followed by a demand peak in the service. This delay in the change of suicide rates or suicidal behaviors has also been observed in other natural disasters [23,24]. In the case of lockdowns associated with the COVID-19 pandemic, the protective effect of pulling together [25] might have also contributed to the reduction of cases.

There is scarcity of studies investigating the impact of COVID-19 on suicidal behaviors providing information on characteristics of cases. Like in our analysis, Yoshioka et al. observed a shift towards younger and female individuals [15]. However, to our knowledge, this is the first analysis of changes in other key features, such as social stressors (e.g., living alone, low socio-economic status, among others) and underlying psychiatric diagnoses. Previous studies in our area found that mental health conditions such as schizophrenia, bipolar disorders, and personality and behavior disorders severely increased the suicide risk [26]. Although complete suicide and suicidal behaviors should be taken as two separate entities [27,28], it is noteworthy that mental health comorbidities were less prevalent among individuals with suicidal behaviors after the COVID-19 onset. Taken together, our results suggest shifting towards lower risk profiles according to classical risk factors; while lower prevalence of living alone and lacking social or family support is consistent with the growth of youngest cases, whether the severity of the risk is actually lower cannot be confirmed due to lack of data on suicide related deaths. Finally, our analysis of individual characteristics before and after the pandemic onset showed that individuals in the low socioeconomic group persistently account for the highest percentage of suicidal behaviors in our area. Although the data should be taken cautiously because, unlike age and gender, which remained constant throughout the analysis period, population distribution across socio-economic groups may have easily changed throughout the period due to economic consequences of the pandemic, our findings suggest a shift towards less economically disadvantaged profiles. This trend is consistent with the incidence analysis. Although the incidence curve seems to increase more abruptly in the low socioeconomic group, the relative increase was similar or even higher in the moderate group (the low socioeconomic group increased from a monthly incidence of 3.96–to–7.92 per 100,000 before the pandemic to 9.05–to–11.59 in the first semester of 2022; the corresponding increase for the moderate socioeconomic group was from 1.35 – 3.36 to 4.09 – 5.43).

Importantly, our findings should be constrained to the setting of suicidal behavior, which may not necessarily reflect trend changes in suicide rate or changes in the characteristics of suicide completers during the pandemic. Yet, since previous suicidal behavior is one of the strongest risk factors for death by suicide [26], our findings raise alarm. Furthermore, some limitations associated with the study design should be considered, such as the uncertainty regarding the quality of reporting, typically seen in retrospective analyses. This is particularly relevant for the criteria of suicidal behavior and referral to the psychiatry ward following the activation of the Suicide Risk Code. Although the Suicide Risk Code program provides physicians with referral instructions [16], establishing suicidal behaviors may not be straightforward in some cases and we cannot rule out heterogeneity between centers regarding these criteria. Finally, underreporting due to overburdening of healthcare centers during the most severe waves of the outbreak, or the effects of a perceived lower accessibility to healthcare services by the population leading to not seeking medical help, should not be ruled out [29]. Despite these limitations, our analysis is strengthened by the population-based approach and the nationwide deployment of the Suicide Risk Code, which allowed the collection of important information about the episode and the individual state, including the suicidality module of the MINI scale.

## 5. Conclusions

Our results raise alarm of the increasing trend of suicidal behaviors within the years following the onset of the COVID-19 pandemic, which continued growing by the end of the investigated period. This increase is primarily explained by a higher incidence of suicidal behaviors among young women. Although we do not know whether these findings are also translated to suicide deaths in our area, actions should be taken to revert this trend. It is worth highlighting that, owing to the younger profile of individuals committing suicide, the potential years of life lost due to suicide may overtake those due to COVID-19 [11]. The notable gender and socioeconomic effects and the shift in the clinical profile of individuals with suicidal behaviors should be taken into account when designing and implementing policies for preventing suicide in the pandemic or post-pandemic context.

## Data Availability

All data produced in the present study are available upon reasonable request to the authors.

## 6. Disclosures

### Supplementary Materials

The following supporting information can be downloaded at: www.mdpi.com/xxx/s1, Table S1: Codes of the International Classification of Diseases (v10, clinical modification) considered for each mental health disorder; Table S2: Type of drug abuse of individuals with suicidal behaviors within the investigated period. Results are presented as no. and percentage of individuals; Figure S1: Decomposition of additive time series; Figure S2. Monthly incidence, overall and according to gender; sub-analysis for individuals within the high (a), moderate (b), and low or very low (c) socio-economic status; Figure S3: Type of drug abuse among individuals with suicidal behaviors. Percentages for the pre-pandemic and pandemic periods..

### Author Contributions

Conceptualization, DV-B, MF, GC-S, and DP; methodology, DV, MF, GC-S, IP, JP, CP, and DP; formal analysis, DV-B; investigation, DV, MF, GC-S, IP, JP, CP, and DP; resources, JP, CP, and DP; data curation, DV-B; writing—original draft preparation, DV-B, MF, and GC-S; writing—review and editing, DV, MF, GC-S, IP, JP, CP, and DP; project administration, GC-S, JP, CP, and DP; funding acquisition, JP. All authors have read and agreed to the published version of the manuscript.

### Institutional Review Board Statement

The study protocol was approved by the independent ethics committee of the Consortium Corporació Sanitària Parc Taulí of Sabadell, which waived the col-lection of informed consent for secondary use of health data collected during routine care. The study was conducted according to the General Data Protection Regulation 2016/679 on data protection and privacy for all individuals within the European Union and the local regulatory framework regarding data protection.

### Informed Consent Statement

Patient consent was waived due to the retrospective nature of the analysis.

### Data Availability Statement

Data regarding suicidal behaviors is highly sensitive; therefore, raw data used in this analysis cannot be publicly available.

## Acknowledgments

The authors would like to thank Sergi Garcia-Trancoso and Toni Fuentes, from the Catalan Health Service, for their contribution in data mining. DP and IP thanks the support of Spanish Ministry of Science and Innovation / ISCIII/FEDER (PI17/01205 and PI21/01148); the Secretaria d’Universitats i Recerca del Departament d’Economia i Coneixement of the Generalitat de Catalunya (2017 SGR 1412); and the CERCA program to the I3PT; the Instituto de Salud Carlos III and the CIBER of Mental Health (CIBERSAM).

## References

1. Flaxman, S.; Mishra, S.; Gandy, A.; Unwin, H.J.T.; Mellan, T.A.; Coupland, H.; Whittaker, C.; Zhu, H.; Berah, T.; Eaton, J.W.; et al. Estimating the effects of non-pharmaceutical interventions on COVID-19 in Europe. Nature 2020, doi:10.1038/s41586-020-2405-7.

2. Mohamad Marzuki, M.F.; Yaacob, N.A.; bin Yaacob, N.M.; Abu Hassan, M.R.; Ahmad, S.B Usable Mobile App for Community Education on Colorectal Cancer: Development Process and Usability Study. JMIR Hum. Factors 2019, 6, e12103, doi:10.2196/12103.

3. Holmes, E.A.; O’Connor, R.C.; Perry, V.H.; Tracey, I.; Wessely, S.; Arseneault, L.; Ballard, C.; Christensen, H.; Silver, R.C.; Everall, I. Multidisciplinary research priorities for the COVID-19 pandemic: a call for action for mental health science. The Lancet Psychiatry 2020.

4. Yang, Y.; Li, W.; Zhang, Q.; Zhang, L.; Cheung, T.; Xiang, Y.-T. Mental health services for older adults in China during the COVID-19 outbreak. The Lancet Psychiatry 2020, 7, e19.

5. Arango, C. Lessons Learned From the Coronavirus Health Crisis in Madrid, Spain: How COVID-19 Has Changed Our Lives in the Last 2 Weeks. Biol. Psychiatry 2020.

6. Arango, C. Lessons Learned From the Coronavirus Health Crisis in Madrid, Spain: How COVID-19 Has Changed Our Lives in the Last 2 Weeks. Biol. Psychiatry 2020, 88, e33–e34, doi:10.1016/j.biopsych.2020.04.003.

7. Fidel Kinori, S.G.; Carot-Sans, G.; Cuartero, A.; Valero-Bover, D.; Piera-Jiménez, J.; Roma Monfa, R.; Garcia, E.; Pérez Sust, P.; Blanch, J.; Ramos-Quiroga, J.A. Web App for Emotional Management During the COVID-19 Pandemic: Platform Development and Retrospective Analysis of Its Use Throughout Two Waves of the Outbreak in Spain (Preprint). JMIR Form. Res. 2021, doi:10.2196/27402.

8. World Health Organization Suicide Available online: https://www.who.int/news-room/fact-sheets/detail/suicide (Accessed on Jul 9, 2022).

9. World Health Organization Suicide worldwide in 2019 Available online: https://www.who.int/publications/i/item/9789240026643 (Accessed on Jul 9, 2022).

10. Ljung, R.; Grunewald, M.; Sundstrom, A.; Sundbom, L.T.; Zethelius, B. Comparison of years of life lost to 1,565 suicides versus 10,650 COVID-19 deaths in 2020 in Sweden: four times more years of life lost per suicide than per COVID-19 death. Ups. J. Med. Sci. 2022, 127, doi:10.48101/ujms.v127.8533.

11. Merayo-Cano, J.M.; Porras-Segovia, A.; Baca-García, E. COVID-19 impact vs. suicide impact in Spain. Rev. Psiquiatr. Salud Ment. 2022, doi:10.1016/J.RPSM.2022.05.006.

12. van der Burgt, M.C.A.; Mérelle, S.; Beekman, A.T.F.; Gilissen, R. The Impact of COVID-19 on the Suicide Prevention Helpline in The Netherlands. Crisis 2022, doi:10.1027/0227-5910/a000863.

13. Reif-Leonhard, C.; Lemke, D.; Holz, F.; Ahrens, K.F.; Fehr, C.; Steffens, M.; Grube, M.; Freitag, C.M.; Kölzer, S.C.; Schlitt, S.; et al. Changes in the pattern of suicides and suicide attempt admissions in relation to the COVID-19 pandemic. Eur. Arch. Psychiatry Clin. Neurosci. 2022, doi:10.1007/s00406-022-01448-y.

14. Demenech, L.M.; Neiva-Silva, L.; Brignol, S.M.S.; Marcon, S.R.; Lemos, S.M.; Tassitano, R.M.; Dumith, S.C Suicide risk among undergraduate students in Brazil in the periods before and during the COVID-19 pandemic: results of the SABES-Grad national survey. Psychol. Med. 2022, 1–13, doi:10.1017/s0033291722001933.

15. 1. Yoshioka, E.; Hanley, S.J.B.; Sato, Y.; Saijo, Y. Impact of the COVID-19 pandemic on suicide rates in Japan through December 2021: An interrupted time series analysis. Lancet Reg. Heal. - West. Pacific 2022, 24, 100480, doi:10.1016/j.lanwpc.2022.100480.

16. Pérez, V.; Elices, M.; Prat, B.; Vieta, E.; Blanch, J.; Alonso, J.; Pifarré, J.; Mortier, P.; Cebrià, A.I.; Campillo, M.T.; et al. The Catalonia Suicide Risk Code: A secondary prevention program for individuals at risk of suicide. J. Affect. Disord. 2020, 268, 201–205, doi:10.1016/j.jad.2020.03.009.

17. Sheehan, D. V; Lecrubier, Y.; Sheehan, K.H.; Amorim, P.; Janavs, J.; Weiller, E.; Hergueta, T.; Baker, R.; Dunbar, G.C The Mini-International Neuropsychiatric Interview (MINI): the development and validation of a structured diagnostic psychiatric interview for DSM-IV and ICD-10. J. Clin. Psychiatry 1998, 59, 22–33.

18. Government of Catalonia Statistical Institute of Catalonia (Idescat) Available online: https://www.idescat.cat/?lang=en (Accessed on Jul 30, 2022).

19. R Core Team R: A language and environment for statistical com-puting Available online: https://www.r-project.org (Accessed on Dec 20, 2021).

20. Wickham, H. Data analysis. In ggplot2. Use R!; Springer, 2016; pp. 189–201.

21. García-Álvarez, L.; de la Fuente-Tomás, L.; García-Portilla, M.P.; Sáiz, P.A.; Lacasa, C.M.; Dal Santo, F.; González-Blanco, L.; Bobes-Bascarán, M.T.; García, M.V.; Vázquez, C.Á. Early psychological impact of the 2019 coronavirus disease (COVID-19) pandemic and lockdown in a large Spanish sample. J. Glob. Health 2020, 10.

22. Jerónimo, M.Á.; Piñar, S.; Samos, P.; González, A.M.; Bellsolà, M.; Sabaté, A.; León, J.; Aliart, X.; Martín, L.M.; Aceña, R.; et al. Suicidal attempt and suicidal ideation during the COVID-19 pandemic compared to previous years. Rev. Psiquiatr. Salud Ment. 2021, doi:10.1016/j.rpsm.2021.11.004.

23. Madianos, M.G.; Evi, K. Trauma and natural disaster: The case of earthquakes in Greece. J. Loss Trauma 2010, 15, 138–150.

24. Orui, M.; Sato, Y.; Tazaki, K.; Kawamura, I.; Harada, S.; Hayashi, M. Delayed increase in male suicide rates in tsunami disaster-stricken areas following the great east Japan earthquake: A three-year follow-up study in Miyagi prefecture. Tohoku J. Exp. Med. 2015, 235, 215–222, doi:10.1620/tjem.235.215.

25. Gordon, K.H.; Bresin, K.; Dombeck, J.; Routledge, C.; Wonderlich, J.A The impact of the 2009 Red River Flood on interpersonal risk factors for suicide. Cris. J. Cris. Interv. Suicide Prev. 2011, 32, 52.

26. Fradera, M.; Ouchi, D.; Prat, O.; Morros, R.; Martin-Fumadó, C.; Palao, D.; Cardoner, N.; Campillo, M.T.; Pérez-Solà, V.; Pontes, C. Can routine Primary Care Records Help in Detecting Suicide Risk? A Population-Based Case-Control Study in Barcelona. Arch. Suicide Res. 2021, 0, 1–15, doi:10.1080/13811118.2021.1911894.

27. Parra Uribe, I.; Blasco-Fontecilla, H.; García-Parés, G.; Giró Batalla, M.; Llorens Capdevila, M.; Cebrià Meca, A.; De Leon-Martinez, V.; Pérez-Solà, V.; Palao Vidal, D.J. Attempted and completed suicide: Not what we expected? J. Affect. Disord. 2013, 150, 840–846, doi:10.1016/j.jad.2013.03.013.

28. Klonsky, E.D.; May, A.M.; Saffer, B.Y Suicide, Suicide Attempts, and Suicidal Ideation. Annu. Rev. Clin. Psychol. 2016, 12, 307–330, doi:10.1146/annurev-clinpsy-021815-093204.

29. Roso-Llorach, A.; Serra-Picamal, X.; Cos, F.X.; Pallejà-Millán, M.; Mateu, L.; Rosell, A.; Almirante, B.; Ferrer, J.; Gasa, M.; Gudiol, C.; et al. Evolving mortality and clinical outcomes of hospitalized subjects during successive COVID-19 waves in Catalonia, Spain. Glob. Epidemiol. 2022, 4, 100071, 1. doi:10.1016/j.gloepi.2022.100071.

