## Supplementary File 1 for "Impact of COVID-19 pandemic on the incidence of suicidal behaviors: a retrospective analysis of integrated electronic health records in a 7.5-million population"

### SUPPLEMENTARY METHODS

Table S1. Codes of the International Classification of Diseases (v10, clinical modification) considered for each mental health disorder

| | |
|---|---|
| F1* | Drugs |
| F10* | Alcohol abuse |
| F11* | Opioids abuse |
| F12* | Cannabis abuse |
| F13* | Hypnotic drugs abuse |
| F14* | Cocaine abuse |
| F17* | Tobacco abuse |
| F51* | Sleep disorders |
| F6* | Personality disorders |
| F31* | Bipolar disorders |
| F33* - F39* | Depression |
| F41* | Anxiety |
| F50* | Eating disorders |
| F20*, F25* | Schizophrenia |
| G30* | Alzheimer |

#### SUPPLEMENTARY FIGURES

Figure S1. Decomposition of additive time series.

The observed rate was decomposed into a time series including the overall trend, the additive seasonal component, and random component using moving averages. The moving average considered a 12-month symmetric window with equal weights. The seasonal component was computed by averaging, for each time unit, over all periods. Finally, the error component is determined by removing trend and seasonal components from the original time series.

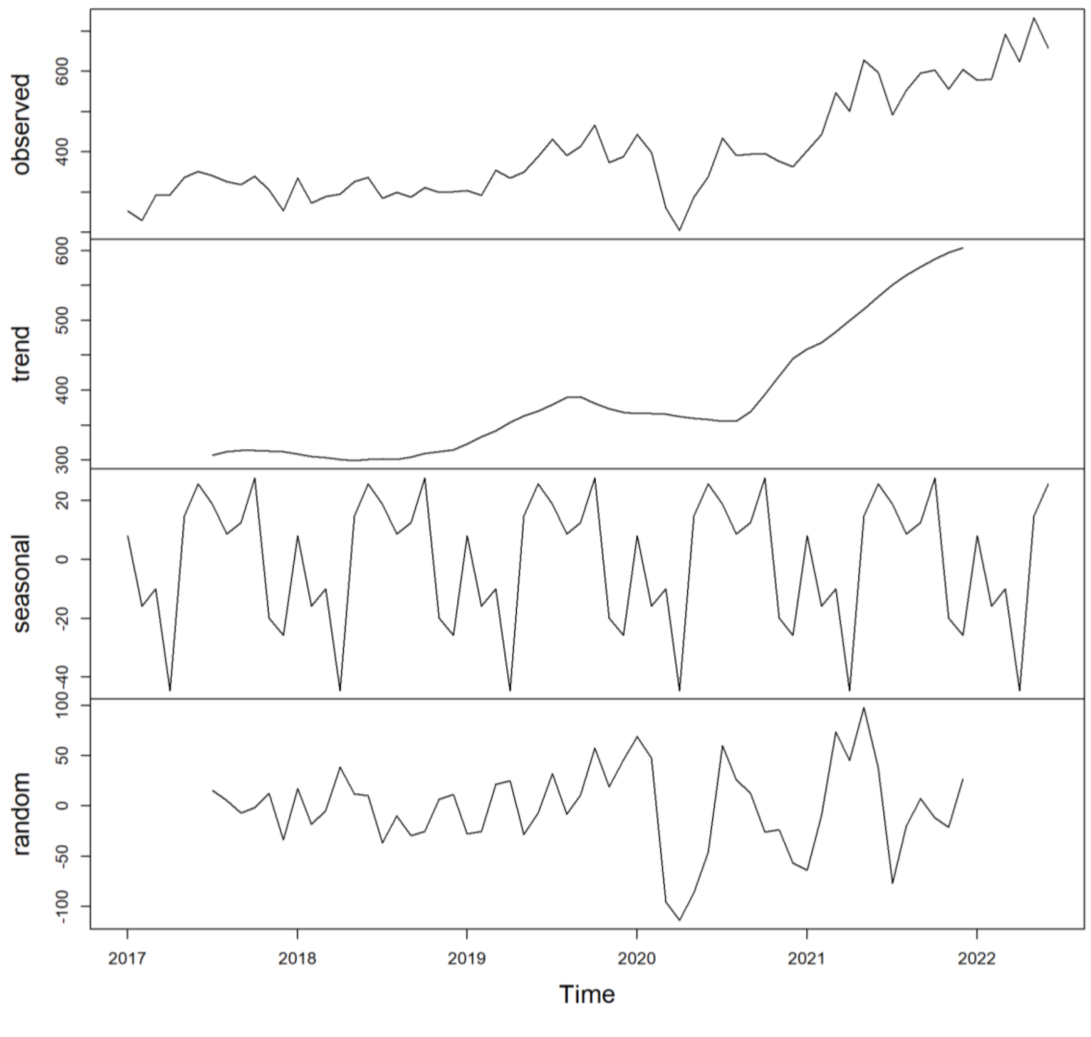

Figure S2. Monthly incidence, overall and according to gender; sub-analysis for individuals within the high (a), moderate (b), and low or very low (c) socioeconomic status.

**a)**

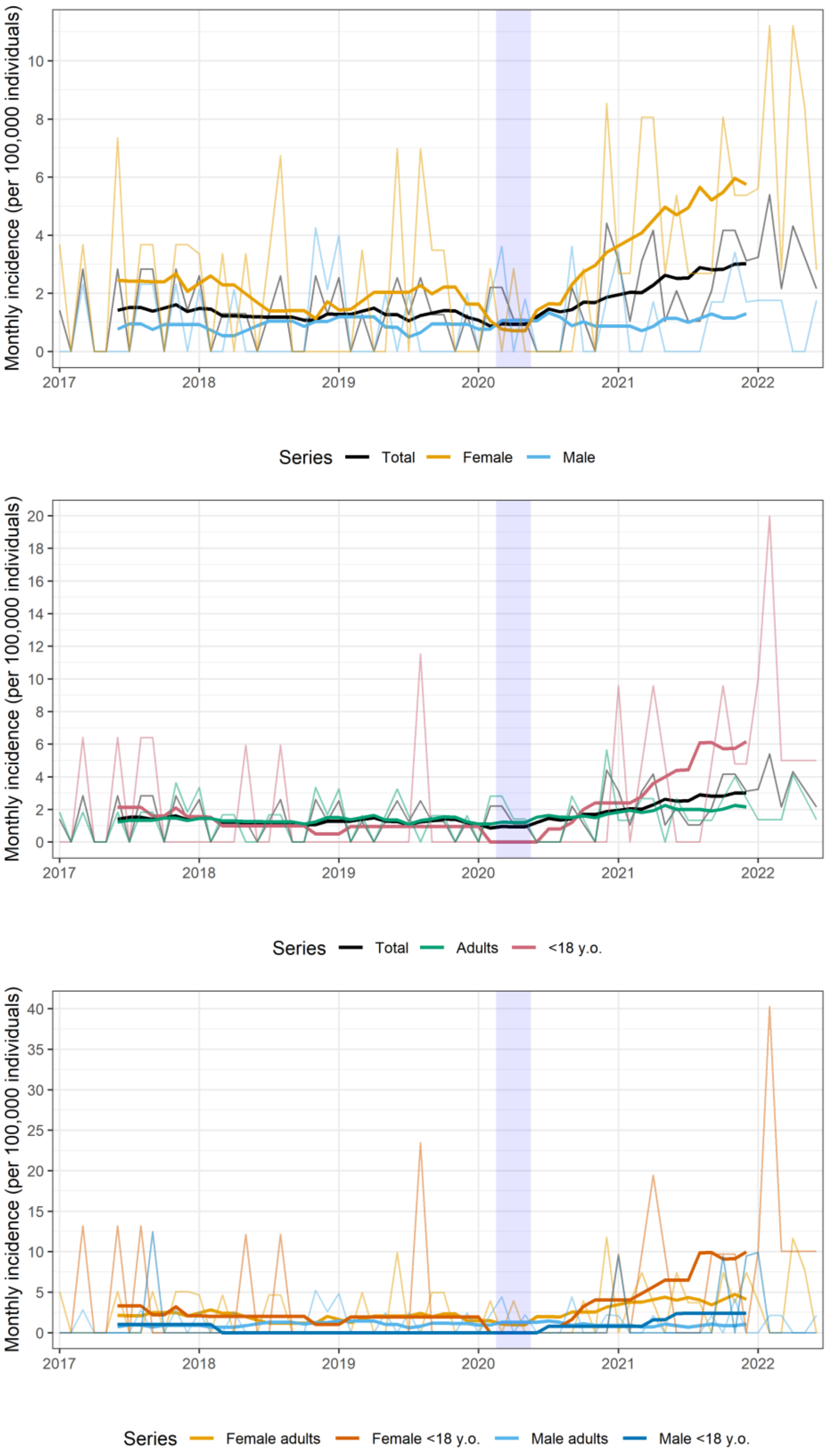

**b)**

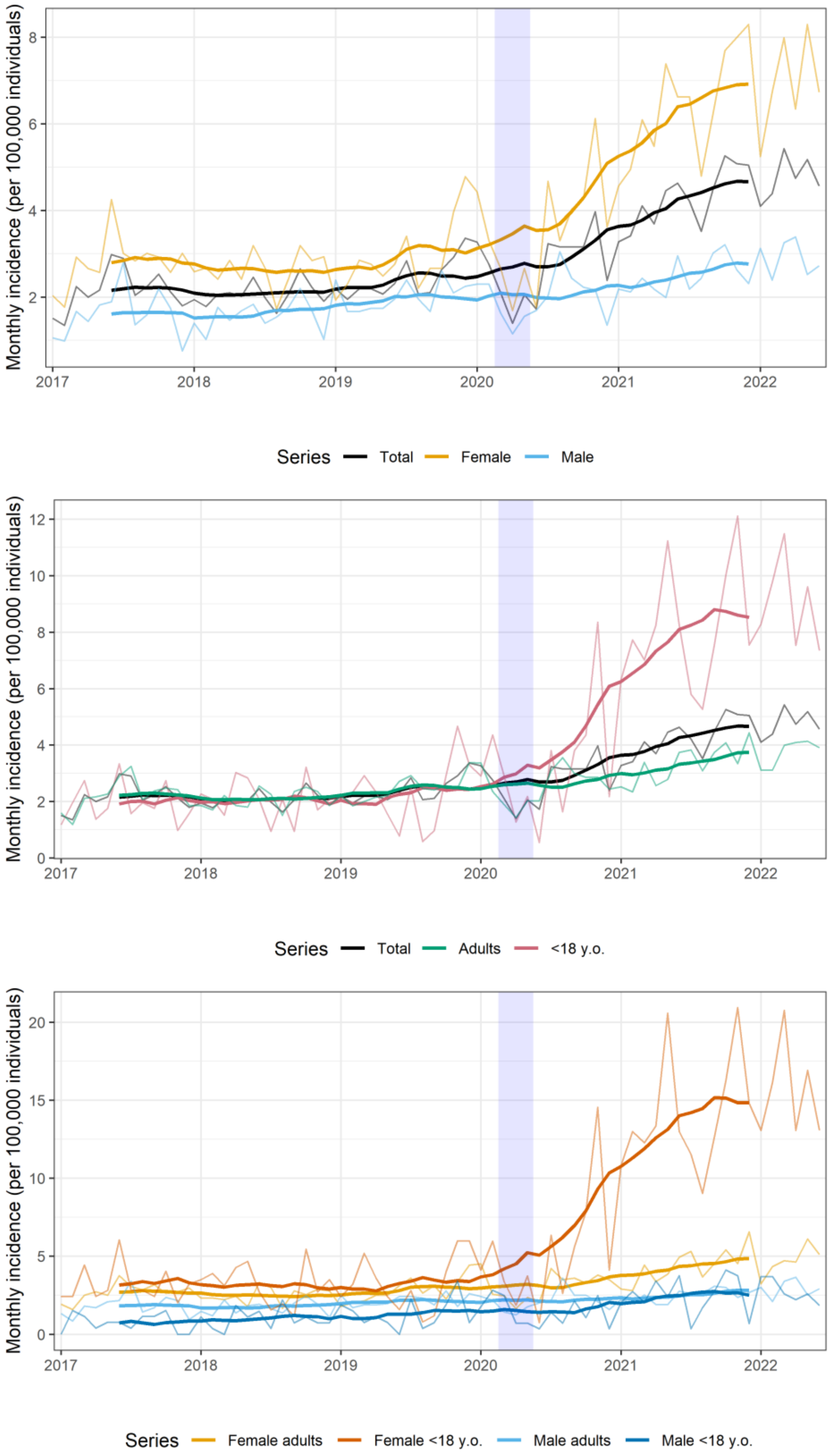
Monthly incidence (per 100,000 individuals)
2017
2018
2019
2020
2021
2022
Series
Total
Female
Male
Adults
<18 y.o.
Female adults
Female <18 y.o.
Male adults
Male <18 y.o.

c)

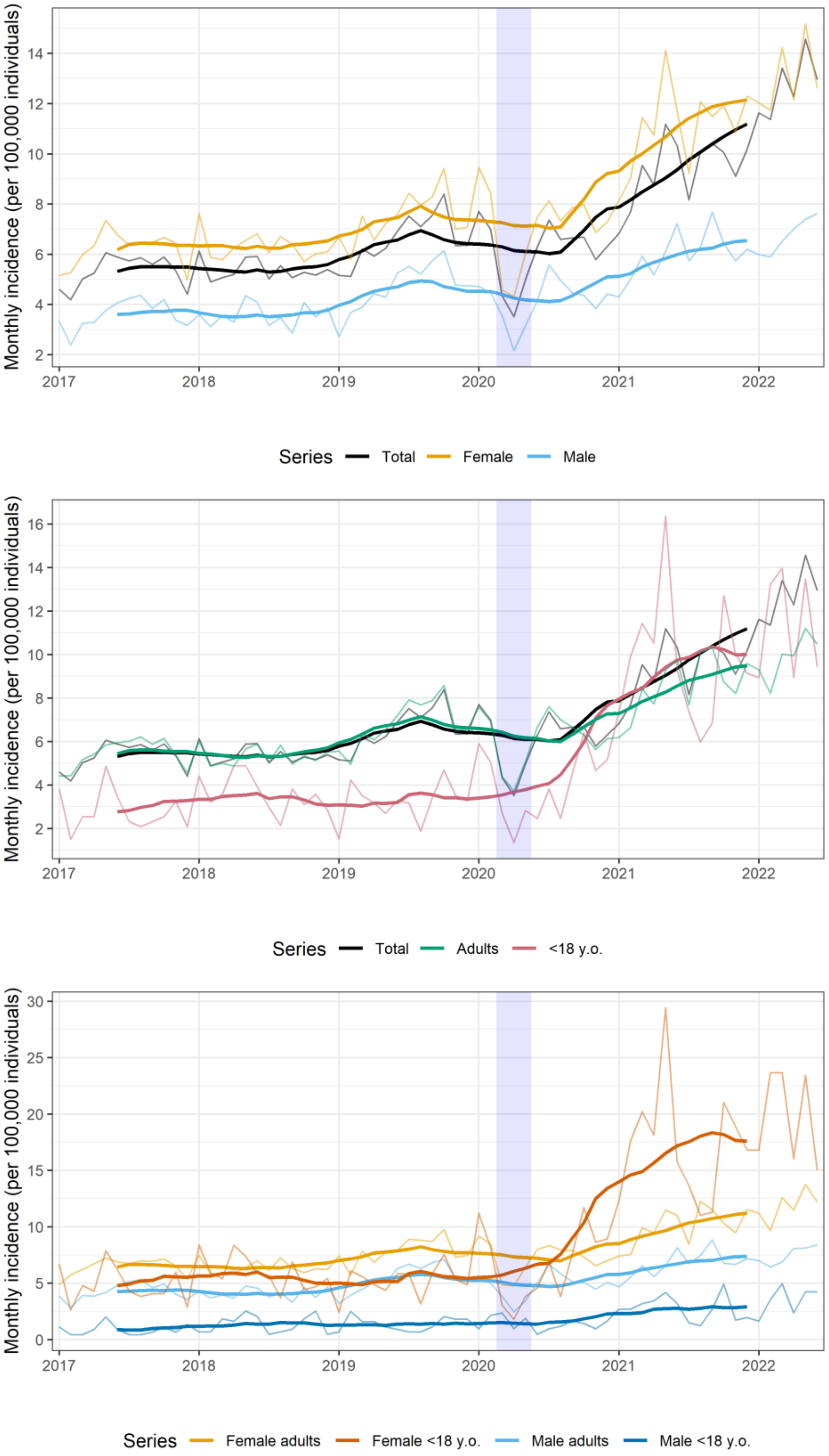
Monthly incidence (per 100,000 individuals)
2
4
6
8
10
12
14
2017
2018
2019
2020
2021
2022
Series
Total
Female
Male
Monthly incidence (per 100,000 individuals)
2
4
6
8
10
12
14
16
2017
2018
2019
2020
2021
2022
Series
Total
Adults
<18 y.o.
Monthly incidence (per 100,000 individuals)
0
5
10
15
20
25
30
2017
2018
2019
2020
2021
2022
Series
Female adults
Female <18 y.o.
Male adults
Male <18 y.o.

Figure S3. Type of drug abuse among individuals with suicidal behaviors. Percentages for the pre-pandemic period (i.e., January 01, 2017 to February 29, 2020) and pandemic period (i.e., March 01, 2020 to June 30, 2022).

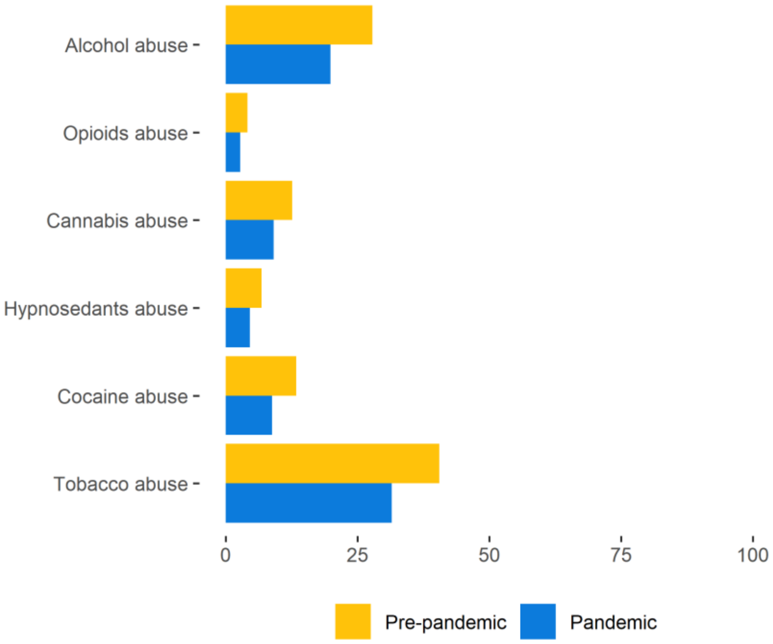

#### SUPPLEMENTARY TABLES

Table S2. Type of drug abuse of individuals with suicidal behaviors within the investigated period. Results are presented as no. and percentage of individuals.

| | **2017** (N = 3641) | **2018** (N = 3638) | **2019** (N = 4489) | **2020** (N = 4291) | **2021** (N = 6528) | **2022**[1] (N = 3871) |
|---|---|---|---|---|---|---|
| Alcohol abuse | 874 (24%) | 773 (21.25%) | 1033 (23.01%) | 1002 (23.35%) | 1149 (17.6%) | 594 (15.34%) |
| Opioids abuse | 132 (3.63%) | 122 (3.35%) | 160 (3.56%) | 152 (3.54%) | 163 (2.5%) | 70 (1.81%) |
| Cannabis abuse | 370 (10.16%) | 346 (9.51%) | 454 (10.11%) | 456 (10.63%) | 548 (8.39%) | 265 (6.85%) |
| Hypnotic drugs abuse | 180 (4.94%) | 187 (5.14%) | 258 (5.75%) | 244 (5.69%) | 275 (4.21%) | 128 (3.31%) |
| Cocaine abuse | 396 (10.88%) | 393 (10.8%) | 536 (11.94%) | 430 (10.02%) | 531 (8.13%) | 272 (7.03%) |
| Tobacco abuse | 1500 (41.2%) | 1464 (40.24%) | 1825 (40.65%) | 1671 (38.94%) | 1976 (30.27%) | 1046 (27.02%) |

[1] The observation period for year 2022 ends on June 30, 2022.
